# Associations of Circulating Biomarkers with Disease Risks: a Two-Sample Mendelian Randomization Study

**DOI:** 10.1101/2024.06.30.24309729

**Authors:** Abdulkadir Elmas, Kevin Spehar, Ron Do, Joseph M. Castellano, Kuan-lin Huang

## Abstract

**Background:** Circulating biomarkers play a pivotal role in personalized medicine, offering potential for disease screening, prevention, and treatment. Despite established associations between numerous biomarkers and diseases, elucidating their causal relationships is challenging. Mendelian Randomization (MR) can address this issue by employing genetic instruments to discern causal links. Additionally, using multiple MR methods with overlapping results enhances the reliability of discovered relationships.

**Methods:** Here we report an MR study using multiple methods, including inverse variance weighted, simple mode, weighted mode, weighted median, and MR Egger. We use the MR-base resource (v0.5.6)^1^ to evaluate causal relationships between 212 circulating biomarkers (curated from UK Biobank analyses by Neale lab and from Shin et al. 2014, Roederer et al. 2015, and Kettunen et al. 2016)^2-4^ and 99 complex diseases (curated from several consortia by MRC IEU and Biobank Japan).

**Results:** We report novel causal relationships found by 4 or more MR methods between glucose and bipolar disorder (Mean Effect Size estimate across methods: 0.39) and between cystatin C and bipolar disorder (Mean Effect Size: -0.31). Based on agreement in 4 or more methods, we also identify previously known links between urate with gout and creatine with chronic kidney disease, as well as biomarkers that may be causal of cardiovascular conditions: apolipoprotein B, cholesterol, LDL, lipoprotein A, and triglycerides in coronary heart disease, as well as lipoprotein A, LDL, cholesterol, and apolipoprotein B in myocardial infarction.

**Conclusions:** This Mendelian Randomization study not only corroborates known causal relationships between circulating biomarkers and diseases but also uncovers two novel biomarkers associated with bipolar disorder that warrant further investigation. Our findings provide insight into understanding how biological processes reflecting circulating biomarkers and their associated effects may contribute to disease etiology, which can eventually help improve precision diagnostics and intervention.

## 1. Introduction

Molecular abnormalities detected in the blood that cause complex diseases represent an opportunity to identify biomarkers for both preventive and therapeutic interventions. These circulating biomolecules include proteins (enzymes and hormones), lipids, and metabolites that reflect physiological states of organ functions, immune response, and metabolism. Despite substantial observational evidence linking systemic biomarker levels with diverse health conditions, their causal relationships to complex diseases remain to be established, especially for diseases for which no reliable biomarkers exist. Previous studies that identify correlations often do not delineate cause and effect, thereby limiting the translational value of the findings. For example, while an increased incidence of impaired glucose metabolism has been demonstrated in patients with bipolar disorder across multiple studies, a causal relationship between the two has yet to be established.^5-7^

The application of Mendelian Randomization (MR) represents an approach in addressing this critical gap. By utilizing genetic instruments as proxies for biomarker levels, MR, under specific assumptions, can control for confounding factors and reverse causation, offering insights into the causal effects of biomarkers on disease risks. Lipids have been particularly well studied in MR, where previous studies have demonstrated the causal relationships between LDL-c and coronary artery disease, and HDL-c and breast cancer.^8-11^ Other studies have also identified likely causal relationships between homocysteine and stroke,^12^ metabolic syndrome,^13^ as well as tyrosine and Type 2 Diabetes.^14^ However, the effects of many other biomarkers remain unclear, and existing MR studies have often been limited to specific biomarkers or disease categories.

The power of MR has been boosted by the availability of large genome-wide association studies (GWAS), published in more than 379 studies including those conducted using the UK Biobank dataset that has recently released a resource of plasma biomarker data measured by nuclear magnetic resonance (NMR) in addition to other lab markers.^15^ Herein, we conducted a comprehensive MR analysis encompassing a broad spectrum of 212 biomarkers—including 115 circulating biomolecules measured through NMR^4^—and 99 human diseases to unveil previously obscured relationships of the circulating biomarkers’ role in disease etiology that may be leveraged for personalized prevention.

## 2. Results

Based on the final 212 exposures (circulating biomarkers) and 99 outcomes (diseases) from the MRC IEU OpenGWAS database^1,16^ (Methods, STROBE-MR checklist), we analyzed exposure-outcome relationships using 5 different MR analysis methods: inverse variance weighted, MR Egger, simple mode, weighted median, and weighted mode. To ensure our results were robust, we focused on findings that had a significant (Bonferroni p-value < 0.05) association by MR-Egger and matched in 4 or more methods (with same effect direction and raw p-value < 0.05) based on MR-Egger’s robustness to directional pleiotropy relevant to circulating biomarkers. We identified a total 21 significant biomarker exposure vs. disease outcome associations (Figure 1). A list of all significant relations, including IEU GWAS study IDs for each exposure/outcomes, and the summary statistics from the MR analyses are included in Supplementary Table 1.

**Figure 1.**
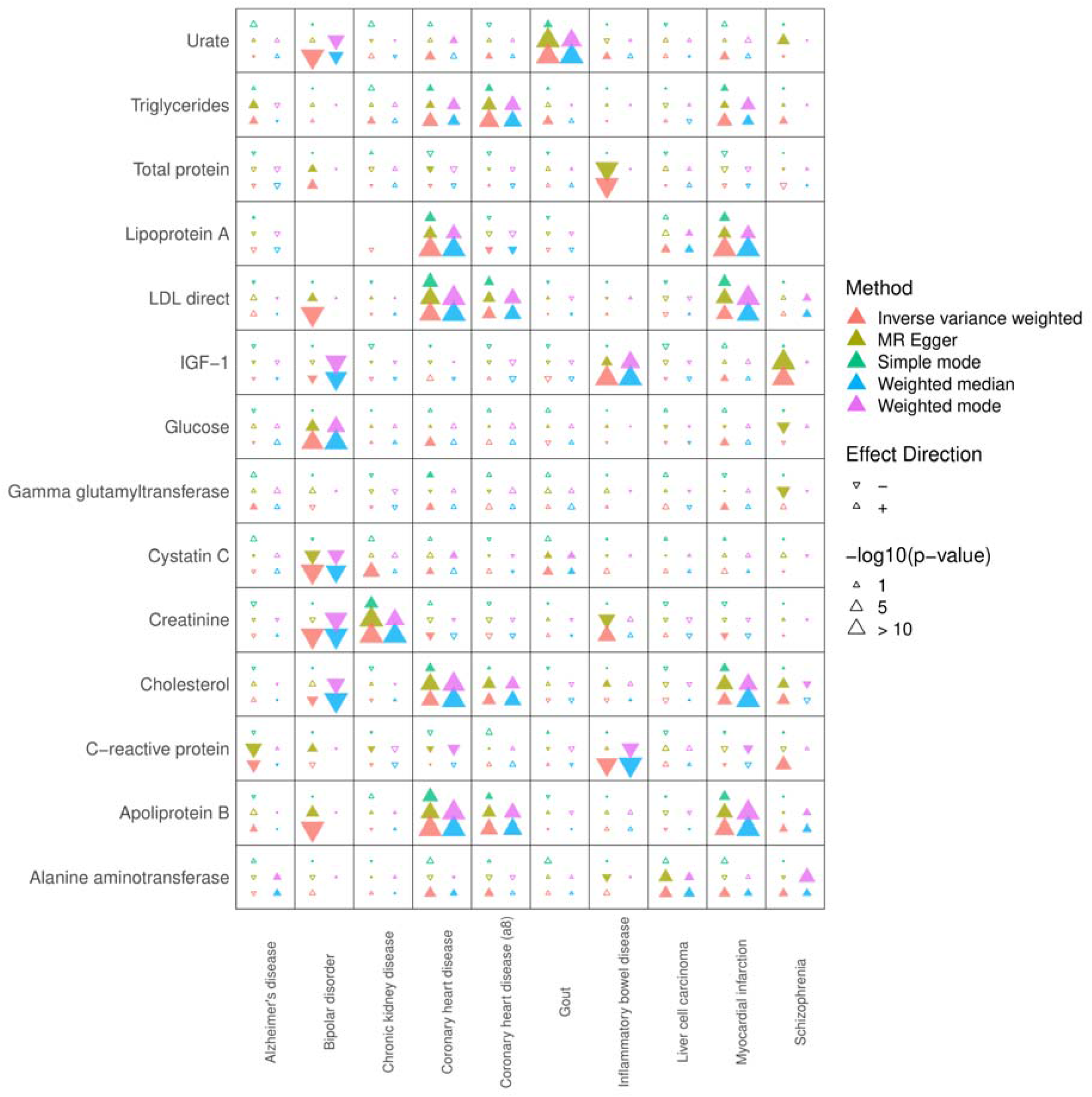
Heatmap summarizing the key findings of this study, including the most significant findings from the MR Egger analysis. MR analyses were conducted using 5 methods: Inverse variance weighted, MR Egger, Simple mode, Weighted Median, and Weighted Mode. Larger arrow sizes correspond to more significant results and the directionality of the arrow indicates a positively-correlated relationship, i.e., higher biomarkers associated with increased risk (upward arrow) or inverse (downward arrow) relationship. We displayed all exposures and outcomes that demonstrated at least one significant (Bonferroni p-value < 0.05) relationship by at least one MR method.

We provide validation for our analysis by demonstrating agreement using 4 or more analysis methods that have been previously demonstrated in MR studies such as urate with gout^17^ and LDL,^18^ triglycerides,^10^ lipoprotein A,^19^ and apolipoprotein B^20^ with coronary heart disease. We also found previously demonstrated relationships shown in MR studies between lipoprotein A^21^ and LDL^22^ with myocardial infarction. Additionally, we did not find any relationships that showed conflicting directionality to the previous literature. While many identified relationships are only maintained in a few methods, previously supported relationships show strong alignment in multiple MR analysis methods, demonstrating the robustness of associations found using multiple approaches.

For cardiometabolic traits, we demonstrated a causal relationship between total cholesterol and coronary heart disease, and this association has previously been shown in a meta-analysis and systematic review.^23^ We also showed a causal relationship between total cholesterol and myocardial infarction as well as apolipoprotein B and myocardial infarction, both of which has been previously demonstrated by non-MR studies.^24,25^

We also found a causal relationship between serum creatinine and chronic kidney disease (CKD). However, serum creatinine is an important biomarker for measuring kidney function, as it is used to clinically estimate glomerular filtration rate to help diagnose CKD.^26,27^ While this connection could explain this result, there may be creatinine-specific effects that may cause or exacerbate CKD which remain to be characterized. We also identified a trend for alanine transferase with liver cell carcinoma that agreed for 4+ methods when using raw p-value cut-offs of < 0.05.

Of note, we discovered a strong, direct relationship between glucose and bipolar disorder and a strong, inverse relationship between cystatin C levels and bipolar disorder which, to the best of our knowledge, has never been directly reported before. Previous studies have demonstrated an association between impaired glucose metabolism and bipolar disorder in addition to increased prevalence of pre-diabetes and type 2 diabetes mellitus.^7,28,29^ Table 1 lists the identified exposure-outcome relationships along with previous supporting literature and evidence type.

**Table 1.** Significant exposure-outcomes relationships consistently identified across 4 or more MR methods. Previous literature demonstrating these relationships and the type of evidence supporting each relationship are also detailed. In evidence type, retrospective, prospective, and cross-sectional indicates cohort studies that demonstrate associations of the biomarker with the disease outcome, whereas meta-analysis and systematic review represent syntheses of such studies that do not suggest causality.

| Exposure | Outcome | Previous Evidence | Evidence Type |
| --- | --- | --- | --- |
| LDL | Coronary Heart Disease | 18 | MR |
| Triglycerides | Coronary Heart Disease | 10 | MR |
| Lipoprotein A | Coronary Heart Disease | 19 | MR |
| Apolipoprotein B | Coronary Heart Disease | 20 | MR |
| Cholesterol | Coronary Heart Disease | 23,30 | Meta-Analysis, Systematic Review, MR |
| Urate | Gout | 17 | MR |
| Lipoprotein A | Myocardial Infarction | 21 | MR |
| LDL | Myocardial Infarction | 22 | MR |
| Cholesterol | Myocardial Infarction | 24 | Retrospective |
| Apolipoprotein B | Myocardial Infarction | 25,31 | Prospective, MR |
| Serum Creatinine | Chronic Kidney Disease | 26,27 | MR, Meta-Analysis |
| Glucose | Bipolar Disorder | 28,29 | Cross Sectional, Meta-Analysis, Systematic Review |
| Cystatin C | Bipolar Disorder | N/A | N/A |

Figure 2 illustrates the top-16 significant (Bonferroni p-value < 0.05) biomarker exposure-disease outcome associations with agreement in effect directionality in 4 or more methods (effect size > |0.1|).

**Figure 2.**
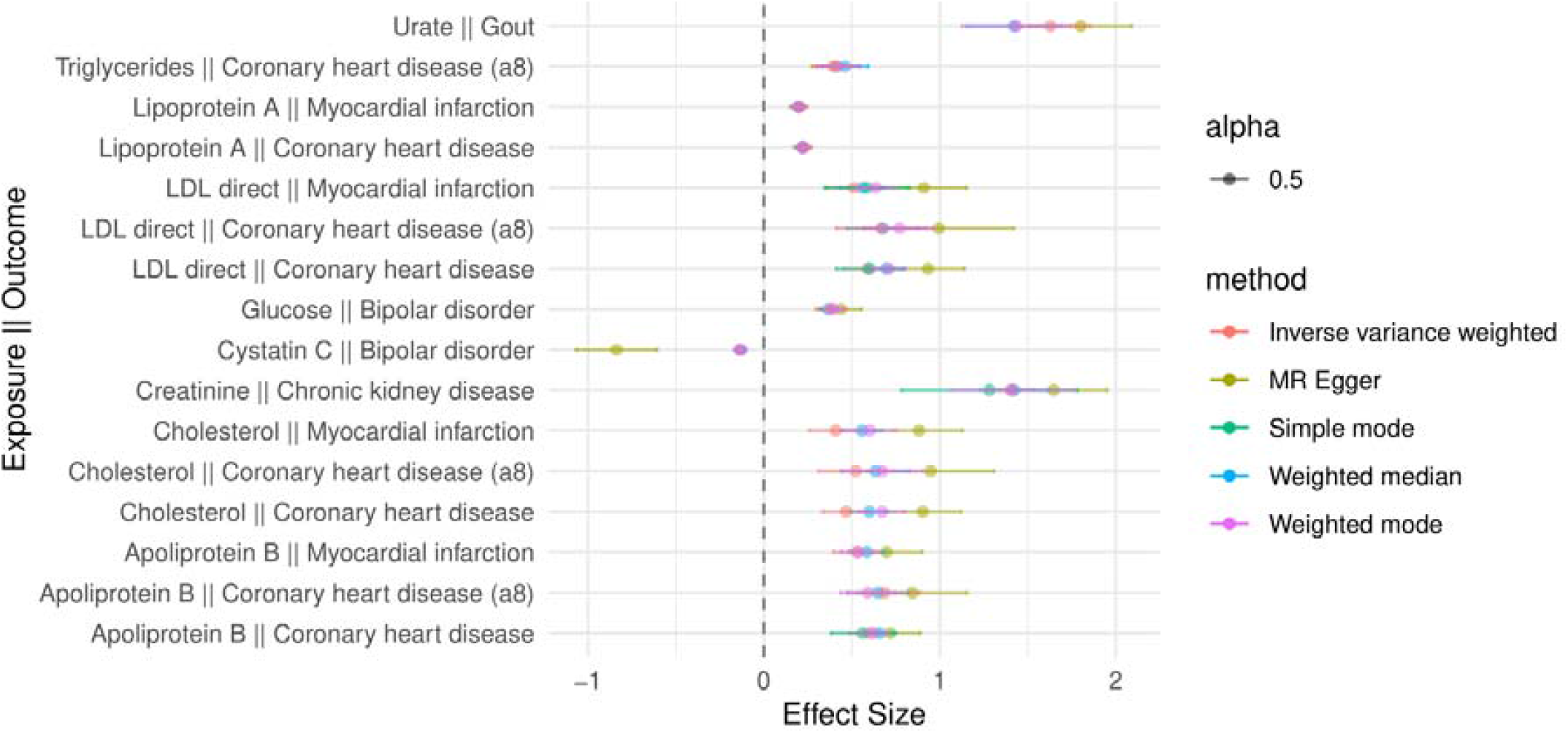
Forest Plot of Mendelian Randomization Analysis Results Assessing the Causal Effects of Circulating Biomarkers on Diseases. The forest plot illustrates the estimated causal effects of top-16 significantly associated (Bonferroni p-value < 0.05) biomarkers on the risk of various diseases based on Mendelian randomization (MR) analysis consistent across 4 or more methods. Each row represents a different MR association, with the biomarker as the exposure and the disease as the outcome. The points represent different effect size estimates from different MR methods and the 95% confidence intervals (based on standard errors of the effect size estimates) are displayed by the horizontal lines forming around the average effect sizes of different methods.

Several biomarkers are associated with an increased risk of disease. The average effect size of total cholesterol and coronary heart disease is 0.66, ranging from 0.90 ± 0.22 (MR Egger) to 0.47 ± 0.13 (IVW). The association of total cholesterol and myocardial infarction has the average effect size of 0.61 with a range of (0.26, 1.13). For the apolipoprotein B and myocardial infarction, the average effect size is estimated as 0.59 with a range of (0.39, 0.90). For the glucose on bipolar disorder the average effect estimate is 0.39 with a range of (0.29, 0.55), while cystatin C has an inverse relationship on bipolar disorder with the average effect size of -0.31 ranging from -0.107 to -1 (Supplementary Table 1).

## 3. Methods

### 3.1. Datasets

This study is a two-sample MR study based on publicly available GWAS summary statistics data from MR-base resource (v0.5.6).^1^ The study design follows the Strengthening the Reporting of Observational Studies in Epidemiology using Mendelian Randomisation (STROBE-MR) checklist, which is included in Supplementary information.

The MR-base platform, developed by the MRC Integrative Epidemiology Unit at the University of Bristol, serves both as a database and an analytical framework accessed through the TwoSampleMR R package (v0.5.7). MR-base offers access to multiple GWAS data including the MRC Integrative Epidemiology Unit (IEU) OpenGWAS database consisting of 26k GWAS reported in at least 379 different studies, covering ∼40k individuals (in median) per study.^1,16^ Based on MRbase, we utilized all accessible exposure traits grouped under “Metabolites” category as well as other circulating biomarkers, and all available outcomes categorized as “Disease”. All diseases we used are from datasets with IDs beginning with “ieu” (GWAS summary datasets generated by many consortia curated by IEU) and “bbj” (Biobank Japan), and the biomarkers are from “ukb” (UK Biobank analyses by IEU or Neale lab covering 28 biomarkers) and “met” (human blood biomarkers, immune markers, and circulating biomarkers analyzed by Shin et al. 2014 covering 76 biomarkers and Kettunen et al. 2016 covering 115 biomarkers).^2,4^ In instances where there are multiple summary datasets for the same trait (i.e., from different GWAS cohorts), we selected the top one from them based on the largest “sample size,” “year,” “number of SNPs,” or “number of cases and controls,” prioritizing the information available in this specified order. A list of all biomarkers used as exposures and information for their corresponding GWAS studies are available in Supplementary Table 2. Overall, we retained a total of 212 exposures (biomarkers) and 99 outcomes (diseases) for MR analyses.

Given that this is a two-sample MR study, we provide two major justifications: (1) Similarity of the Genetic Variant-Exposure Associations Between the Exposure and Outcome Samples: Genetic instruments for the exposure were primarily derived from GWAS of circulating biomarkers conducted using data from the UK Biobank in addition to Shin and Kettunen et al., all of which predominantly consists of individuals of European ancestry. The outcome data were mainly sourced from case-control GWAS of European populations that likely share genetic architectures similar to those of the exposure study participants. (2) Number of Individuals Who Overlap Between the Exposure and Outcome Studies: Given the separate sources of the genetic data for exposures and outcomes (various case-control studies), and the timing of data availability and publication, it is reasonable to assume minimal overlap in the individual participants across these studies. Particularly, all significant results were found based on exposure (biomarker) GWAS from UK Biobank, which has its first genetic data published in 2018. The significant outcome GWAS for bipolar disorder, cardiovascular disease (CVD), myocardial infarction (MI), gout, and chronic kidney disease, all predated 2018. This temporal discrepancy between the data collection phases further supports the assumption of negligible overlap bias in our MR estimates.

### 3.2. MR Methods

The three core assumptions for Mendelian Randomization (MR) analysis are:

#### Relevance

The genetic instruments (e.g., single nucleotide polymorphisms, SNPs) used as instrumental variables (IVs) should be associated with the exposure (circulating biomarkers in this case). This assumption ensures that the IVs are strong enough to influence the exposure. Herein, 2685 SNPs were found associated with 198 circulating biomarker exposures subject to the IV assumptions that were imposed by the standard two-sample MR approach (association threshold, P<5e-8; LD clumping cutoff r2>0.001 within 10Mb window) and were used for further analyses (Supplementary Table 3).

#### Independence

The genetic instruments should be independent of any confounding factors that may affect both the exposure and the outcome (disease). This assumption is justified by the random allocation of genetic variants. Further, each of the GWAS studies carefully control for covariates that may confound genetic associations. For example, in the UK Biobank biomarker GWAS conducted by the Neale lab, covariates include age, sex, age2, age*sex, age*sex2, and the first 20 PCs, making the final genetic instruments less prone to confounding. Additionally, the prior knowledge of demographic consistency can likely minimize the impact of unmeasured confounders due to similar demographic and genetic backgrounds.

#### Exclusion restriction

The genetic instruments should affect the outcome (disease) only through their effects on the exposure (circulating biomarkers) and not through any other direct or indirect pathways. This assumption ensures that the IVs influence the outcome solely via the exposure of interest. Critically, MR Egger was employed as a primary MR method in this study to detect and adjust for pleiotropy, where genetic variants may influence the outcome via pathways other than through the exposure.

To enhance robustness, we inferred causality using multiple MR methods, specifically inverse variance weighted, simple mode, weighted mode, weighted median, and MR Egger. The inverse variance weighted (IVW) method conducts regression analysis between the SNP-exposure effect size and the SNP-outcome effect size, giving more importance to SNPs with the lowest standard error in SNP-outcome association. The IVW method relies on either all variants being valid instruments or the presence of balanced horizontal pleiotropy, where the combined horizontal pleiotropic effects of individual instruments cancel out. Additionally, it assumes that such pleiotropic effects are not dependent on instrument strength across all variants, a concept known as the Instrument Strength Independent of Direct Effects (InSIDE) assumption.^32^ The simple mode method operates under the assumption that the most common value (i.e., mode) for horizontal pleiotropy is zero, known as the ZEro Modal Pleiotropy Assumption (ZEMPA), regardless of the specific type of horizontal pleiotropy.^33^ Likewise, the weighted MBE method operates under the supposition that the most substantial weights in the k subsets originate from valid instruments, adhering to ZEMPA assumption, irrespective of the particular form of horizontal pleiotropy. The weighted median method relies on the assumption that over half of the weight comes from reliable instruments, irrespective of the nature of horizontal pleiotropy.^34^ MR-Egger permits all genetic variants to exhibit pleiotropic effects, but it necessitates that these effects are independent of the SNP–exposure associations, following the InSIDE principle.^35^ Notably, the method can correct for the average horizontal pleiotropy observed across all variants considered in the study. Considering the recognized pleiotropic effects commonly seen in biomarkers and their relevance to health and disease,^36^ we used MR-Egger as the primary method for determining important exposure-outcome associations in our analyses and employed other sensitivity methods for validation.

For each method, the associations are reported with effect size and corresponding p-value statistics. The raw p-values are multi-testing corrected using the Bonferroni method. The confidence intervals (95%) are also calculated from standard errors of the test statistics. Overall, robustness of the results is ensured based on the use of MR-Egger as discovery method, agreement across at least 4 MR methods, and Bonferroni correction.

## 4. Discussion

We analyzed exposure-outcome relationships using 5 MR analysis methods to identify causal relationships between circulating biomarkers and diseases. In this study, significant results are defined as agreement between 4 or more analysis methods. Many of our demonstrated results replicated results from previous MR studies and other literature and none of our findings run contrary to previous literature. Additionally, our analysis also discovered two relationships of note: a direct relationship between glucose and bipolar disorder and an inverse relationship between cystatin C and bipolar disorder.

Bipolar disorder is a neuropsychiatric disorder with multifactorial causes (genetic, trauma, exposure to certain medications, etc.) Our findings demonstrate two potential circulating markers that may contribute to development of bipolar disorder, high glucose and low cystatin C. In our study, the effect size for glucose and bipolar disorder is 0.39 and for cystatin C and bipolar disorder is -0.31 which is comparable to the average effect size for triglycerides and coronary heart disease (a8) of 0.42 and greater than those of triglycerides and myocardial infarction (0.20), lipoprotein A and coronary heart disease (0.22), and lipoprotein A and myocardial infarction (0.21) (Supplementary Table 1). These results suggest that these relationships are likely to be of clinical significance and warrants further study. Although high cystatin C levels have been linked with major depressive disorder,^37^ no studies, to the best of our knowledge, have demonstrated an association between cystatin C and bipolar disorder. Levels of Cystatin C, a natural inhibitor of cysteine proteases, are typically used clinically to assess kidney function. A common treatment for bipolar disorder is lithium, which has been shown to decrease renal function and can lead to elevated cystatin C levels,^38^ which may lend further insight to the mechanism behind lithium’s use as a treatment, however this remains to be confirmed.

A connection between impaired glucose metabolism and bipolar disorder has been well documented in the literature, with over half of patients diagnosed with bipolar disorder also having insulin resistance, impaired glucose tolerance, or type 2 diabetes.^7^ Furthermore, it has been shown that modulating the PI3K/Akt insulin signaling pathway may be a mechanism of lithium for the treatment of bipolar disorder.^39^ The PI3K/Akt pathway helps regulate glucose metabolism in the hippocampus, cerebellum, and olfactory bulb^40^ which are vulnerable sites of gray matter loss in bipolar disorder.^41^ It has been speculated that long-term disruption of this pathway can lead to mitochondrial dysfunction and energy dysregulation which may affect brain processes contributing to bipolar disorder.^39^ Therefore, our identification of a causal relationship between high glucose levels and bipolar disorder further supports the current body of evidence, and our findings for cystatin C highlight a novel finding that warrants investigation in subsequent studies. Our findings suggest that future studies aiming to reduce the risk of bipolar disorder could explore the lifestyle and clinical interventions that reduce blood glucose and maintain kidney function.

Although our study focused on links that were aligned with 4 or more MR methods, other notable relationships were identified that were aligned with 2-3 MR methods (Figure 1). For example, C-reactive protein (CRP) and Alzheimer’s disease were causally linked in 2 methods (MR Egger and IVW) which has been previously noted in previous non-MR studies.^42^

A limitation of this study is that much of the exposure GWAS data comes from the UK biobank, a large-scale volunteer databank, which has a biased representation of Europeans and results may not generalize outside UK.^43,44^ However, UK biobank provides the largest GWAS datasets of most biomarkers studied herein and adequate statistical power; this bias may be overcome as other biobanks with diverse populations continue to expand. MR studies have their own limitations, including but not limited to: potential confounding factors, limitations of estimating associations for binary outcomes, pleiotropic effects which should be mitigated by our use of MR Egger, and population stratification.^45-47^

Overall, by utilizing several MR methods as cross-validation, our study successfully reaffirmed several established connections in cardiovascular diseases, gout, and kidney diseases, while uncovering intriguing novel biomarkers that may be causal for bipolar disorder. These findings highlight the underappreciated roles that circulating biomarkers may play in disease mechanisms, which should open new avenues for targeted research and motivate development of precise diagnostic tools and therapeutic interventions.

## Supporting information

STROBE-MR checklist

## Data Availability

Data Availability Statement: Data are contained within the article and supplementary materi-als.
Data Availability: GWAS summary statistics data used can be found on MR-base resource (v0.5.6) through the ieu open GWAS project at the URL: https://gwas.mrcieu.ac.uk/.
Code Availability: The source code for the Mendelian randomization analyses is available at github.com/Huang-lab/MR.

## Supplementary Materials

The following supporting information can be downloaded at: www.mdpi.com/xxx/s1, Figure S1: title; Table S1: title; Video S1: title.

## Author Contributions

AE designed and conducted the analyses. KS performed the literature search and interpreted the results. KS, AE, JMC, and KH wrote the manuscript. KH supervised the study. All authors read, edited, and approved the manuscript.

## Institutional Review Board Statement

Not applicable. Our analyses were conducted using publicly available summary statistics data from GWAS, not individual-level data; therefore, this is a non-human subject research that requires no additional ethical approval.

## Data Availability Statement

Data are contained within the article and supplementary materials.

## Data Availability

GWAS summary statistics data used can be found on MR-base resource (v0.5.6) through the ieu open GWAS project at the URL: https://gwas.mrcieu.ac.uk/.

## Code Availability

The source code for the Mendelian randomization analyses is available at github.com/Huang-lab/MR.

## Acknowledgments

The authors wish to acknowledge providers of GWAS summary datasets. The authors thank all members of the Huang lab for constructive discussion. Large language models (LLM) may have been used in the initial drafts of coding and writing of this work. All final codes and texts have been extensively edited and verified by the authors. This work was supported in part through the computational and data resources and staff expertise provided by Scientific Computing and Data at the Icahn School of Medicine at Mount Sinai and supported by the Clinical and Translational Science Awards (CTSA) grant UL1TR004419 from the National Center for Advancing Translational Sciences. Research reported in this publication was also supported by the Office of Research Infrastructure of the National Institutes of Health under award number S10OD026880 and S10OD030463. The content is solely the responsibility of the authors and does not necessarily represent the official views of the National Institutes of Health. This work was supported by NIH NIGMS R35GM138113 and ACS RSG-22-115-01-DMC to KH and RF1AG072300 to JMC.

## Disclaimer/Publisher’s Note

The statements, opinions and data contained in all publications are solely those of the individual author(s) and contributor(s) and not of MDPI and/or the editor(s). MDPI and/or the editor(s) disclaim responsibility for any injury to people or property resulting from any ideas, methods, instructions or products referred to in the content.

## References

1. Hemani G, Zheng J, Elsworth B, et al. The MR-Base platform supports systematic causal inference across the human phenome. Elife 2018;7. DOI:10.7554/eLife.34408.

2. Shin SY, Fauman EB, Petersen AK, et al. An atlas of genetic influences on human blood metabolites. Nat Genet 2014;46(6):543–550. DOI:10.1038/ng.2982.

3. Roederer M, Quaye L, Mangino M, et al. The genetic architecture of the human immune system: a bioresource for autoimmunity and disease pathogenesis. Cell 2015;161(2):387–403. DOI:10.1016/j.cell.2015.02.046.

4. Kettunen J, Demirkan A, Wurtz P, et al. Genome-wide study for circulating metabolites identifies 62 loci and reveals novel systemic effects of LPA. Nat Commun 2016;7:11122. DOI:10.1038/ncomms11122.

5. Ruzickova MS, C. Garnham, J. Alda M. Clinical Features of Bipolar Disorder With and WithoutComorbid Diabetes Mellitus. The Canadian Journal of Psychiatry 2003;48(7):431–502.

6. Calkin CV, Ruzickova M, Uher R, et al. Insulin resistance and outcome in bipolar disorder. Br J Psychiatry 2015;206(1):52–7. DOI:10.1192/bjp.bp.114.152850.

7. Lojko D, Owecki M, Suwalska A. Impaired Glucose Metabolism in Bipolar Patients: The Role of Psychiatrists in Its Detection and Management. Int J Environ Res Public Health 2019;16(7). DOI:10.3390/ijerph16071132.

8. Beeghly-Fadiel A, Khankari NK, Delahanty RJ, et al. A Mendelian randomization analysis of circulating lipid traits and breast cancer risk. Int J Epidemiol 2020;49(4):1117–1131. DOI:10.1093/ije/dyz242.

9. Burgess S, Harshfield E. Mendelian randomization to assess causal effects of blood lipids on coronary heart disease: lessons from the past and applications to the future. Curr Opin Endocrinol Diabetes Obes 2016;23(2):124–30. DOI:10.1097/MED.0000000000000230.

10. Holmes MV, Asselbergs FW, Palmer TM, et al. Mendelian randomization of blood lipids for coronary heart disease. Eur Heart J 2015;36(9):539–50. DOI:10.1093/eurheartj/eht571.

11. Thomas DG, Wei Y, Tall AR. Lipid and metabolic syndrome traits in coronary artery disease: a Mendelian randomization study. J Lipid Res 2021;62:100044. DOI:10.1194/jlr.P120001000.

12. Larsson SC, Traylor M, Markus HS. Homocysteine and small vessel stroke: A mendelian randomization analysis. Ann Neurol 2019;85(4):495–501. DOI:10.1002/ana.25440.

13. Lee HS, In S, Park T. The Homocysteine and Metabolic Syndrome: A Mendelian Randomization Study. Nutrients 2021;13(7). DOI:10.3390/nu13072440.

14. Jager S, Cuadrat R, Wittenbecher C, et al. Mendelian Randomization Study on Amino Acid Metabolism Suggests Tyrosine as Causal Trait for Type 2 Diabetes. Nutrients 2020;12(12). DOI:10.3390/nu12123890.

15. Julkunen H, Cichonska A, Tiainen M, et al. Atlas of plasma NMR biomarkers for health and disease in 118,461 individuals from the UK Biobank. Nat Commun 2023;14(1):604. DOI:10.1038/s41467-023-36231-7.

16. Elsworth B, Lyon M, Alexander T, et al. The MRC IEU OpenGWAS data infrastructure. bioRxiv 2020. DOI:10.1101/2020.08.10.244293.

17. Adams CD, Boutwell BB. Using multiple Mendelian randomization approaches and genetic correlations to understand obesity, urate, and gout. Sci Rep 2021;11(1):17799. DOI:10.1038/s41598-021-97410-4.

18. Ference BA. Mendelian randomization studies: using naturally randomized genetic data to fill evidence gaps. Curr Opin Lipidol 2015;26(6):566–71. DOI:10.1097/MOL.0000000000000247.

19. Wang S, Zha L, Chen J, et al. The relationship between lipoprotein(a) and risk of cardiovascular disease: a Mendelian randomization analysis. Eur J Med Res 2022;27(1):211. DOI:10.1186/s40001-022-00825-6.

20. Richardson TG, Sanderson E, Palmer TM, et al. Evaluating the relationship between circulating lipoprotein lipids and apolipoproteins with risk of coronary heart disease: A multivariable Mendelian randomisation analysis. PLoS Med 2020;17(3):e1003062. DOI:10.1371/journal.pmed.1003062.

21. Nordestgaard BG, Langsted A. Lipoprotein (a) as a cause of cardiovascular disease: insights from epidemiology, genetics, and biology. J Lipid Res 2016;57(11):1953–1975. DOI:10.1194/jlr.R071233.

22. Pham K, Mulugeta A, Lumsden A, Hyppl7lnen E. Genetically instrumented LDL-cholesterol lowering and multiple disease outcomes: A Mendelian randomization phenome-wide association study in the UK Biobank. Br J Clin Pharmacol 2023;89(10):2992–3004. DOI:10.1111/bcp.15793.

23. Peters SA, Singhateh Y, Mackay D, Huxley RR, Woodward M. Total cholesterol as a risk factor for coronary heart disease and stroke in women compared with men: A systematic review and meta-analysis. Atherosclerosis 2016;248:123–31. DOI:10.1016/j.atherosclerosis.2016.03.016.

24. Kim MK, Han K, Kim HS, et al. Cholesterol variability and the risk of mortality, myocardial infarction, and stroke: a nationwide population-based study. Eur Heart J 2017;38(48):3560–3566. DOI:10.1093/eurheartj/ehx585.

25. Walldius G, Jungner I, Holme I, Aastveit AH, Kolar W, Steiner E. High apolipoprotein B, low apolipoprotein A-I, and improvement in the prediction of fatal myocardial infarction (AMORIS study): a prospective study. Lancet 2001;358(9298):2026–33. DOI:10.1016/S0140-6736(01)07098-2.

26. Park S, Lee S, Kim Y, et al. Mendelian randomization reveals causal effects of kidney function on various biochemical parameters. Commun Biol 2022;5(1):713. DOI:10.1038/s42003-022-03659-4.

27. Levey AS, de Jong PE, Coresh J, et al. The definition, classification, and prognosis of chronic kidney disease: a KDIGO Controversies Conference report. Kidney Int 2011;80(1):17–28. DOI:10.1038/ki.2010.483.

28. Mansur RB, Rizzo LB, Santos CM, et al. Impaired glucose metabolism moderates the course of illness in bipolar disorder. J Affect Disord 2016;195:57–62. DOI:10.1016/j.jad.2016.02.002.

29. Vancampfort D, Mitchell AJ, De Hert M, et al. Prevalence and predictors of type 2 diabetes mellitus in people with bipolar disorder: a systematic review and meta-analysis. J Clin Psychiatry 2015;76(11):1490–9. DOI:10.4088/JCP.14r09635.

30. Liao LZ, Li WD, Liu Y, Li JP, Zhuang XD, Liao XX. Exploring the causal pathway from omega-6 levels to coronary heart disease: A network Mendelian randomization study. Nutr Metab Cardiovasc Dis 2020;30(2):233–240. DOI:10.1016/j.numecd.2019.09.013.

31. Ding Y, Yang S, He M, Fan S, Tao X, Lu W. Lipid Metabolism Traits Mediate the Effect of Psoriasis on Myocardial Infarction Risk: A Two-Step Mendelian Randomization Study. Metabolites 2023;13(9). DOI:10.3390/metabo13090976.

32. Burgess S, Butterworth A, Thompson SG. Mendelian randomization analysis with multiple genetic variants using summarized data. Genet Epidemiol 2013;37(7):658–65. DOI:10.1002/gepi.21758.

33. Hartwig FP, Davey Smith G, Bowden J. Robust inference in summary data Mendelian randomization via the zero modal pleiotropy assumption. Int J Epidemiol 2017;46(6):1985–1998. DOI:10.1093/ije/dyx102.

34. Bowden J, Davey Smith G, Haycock PC, Burgess S. Consistent Estimation in Mendelian Randomization with Some Invalid Instruments Using a Weighted Median Estimator. Genet Epidemiol 2016;40(4):304–14. DOI:10.1002/gepi.21965.

35. Bowden J, Davey Smith G, Burgess S. Mendelian randomization with invalid instruments: effect estimation and bias detection through Egger regression. Int J Epidemiol 2015;44(2):512–25. DOI:10.1093/ije/dyv080.

36. Smith CJ, Sinnott-Armstrong N, Cichonska A, et al. Integrative analysis of metabolite GWAS illuminates the molecular basis of pleiotropy and genetic correlation. Elife 2022;11. DOI:10.7554/eLife.79348.

37. Sun T, Chen Q, Li Y. Associations of serum cystatin C with depressive symptoms and suicidal ideation in major depressive disorder. BMC Psychiatry 2021;21(1):576. DOI:10.1186/s12888-021-03509-3.

38. Dastych M, Synek O, Gottwaldova J. Impact of Long-Term Lithium Treatment on Renal Function in Patients With Bipolar Disorder Based on Novel Biomarkers. J Clin Psychopharmacol 2019;39(3):238–242. DOI:10.1097/JCP.0000000000001030.

39. Campbell IH, Campbell H, Smith DJ. Insulin signaling as a therapeutic mechanism of lithium in bipolar disorder. Transl Psychiatry 2022;12(1):350. DOI:10.1038/s41398-022-02122-6.

40. Gabbouj S, Ryhänen S, Marttinen M, et al. Altered Insulin Signaling in Alzheimer’s Disease Brain – Special Emphasis on PI3K-Akt Pathway. Frontiers in Neuroscience 2019;13 (Mini Review) (In English). DOI:10.3389/fnins.2019.00629.

41. Moorhead TWJ, McKirdy J, Sussmann JED, et al. Progressive Gray Matter Loss in Patients with Bipolar Disorder. Biological Psychiatry 2007;62(8):894–900. DOI:10.1016/j.biopsych.2007.03.005.

42. Huang J, Tao Q, Ang TFA, et al. The impact of increasing levels of blood C-reactive protein on the inflammatory loci SPI1 and CD33 in Alzheimer’s disease. Transl Psychiatry 2022;12(1):523. DOI:10.1038/s41398-022-02281-6.

43. Hanlon P, Jani BD, Nicholl B, Lewsey J, McAllister DA, Mair FS. Associations between multimorbidity and adverse health outcomes in UK Biobank and the SAIL Databank: A comparison of longitudinal cohort studies. PLoS Med 2022;19(3):e1003931. DOI:10.1371/journal.pmed.1003931.

44. Brayne C, Moffitt TE. The limitations of large-scale volunteer databases to address inequalities and global challenges in health and aging. Nat Aging 2022;2(9):775–783. DOI:10.1038/s43587-022-00277-x.

45. Glynn RJ. Promises and limitations of mendelian randomization for evaluation of biomarkers. Clin Chem 2010;56(3):388–90. DOI:10.1373/clinchem.2009.142513.

46. Haycock PC, Burgess S, Wade KH, Bowden J, Relton C, Davey Smith G. Best (but oft-forgotten) practices: the design, analysis, and interpretation of Mendelian randomization studies. Am J Clin Nutr 2016;103(4):965–78. DOI:10.3945/ajcn.115.118216.

47. Smith GD, Ebrahim S. Mendelian randomization: prospects, potentials, and limitations. Int J Epidemiol 2004;33(1):30–42. DOI:10.1093/ije/dyh132.

