## Supplementary material for "Associations of Circulating Biomarkers with Disease Risks: a Two-Sample Mendelian Randomization Study": STROBE-MR checklist

**STROBE-MR checklist of recommended items to address in reports of Mendelian randomization studies**^1^ ^2^

| **Item No.** | **Section** | **Checklist item** | **Page No.** | **Relevant text from manuscript** |
| --- | --- | --- | --- | --- |
| 1 | **TITLE and ABSTRACT** | Indicate Mendelian randomization (MR) as the study’s design in the title and/or the abstract if that is a main purpose of the study | 1-2 | Title: Associations of the Circulating Biomarkers with Disease Risks: a Two-Sample Mendelian Randomization Study  Abstract: Here we report an MR study using multiple methods, including inverse variance weighted, simple mode, weighted mode, weighted median, and MR Egger. |
|  | **INTRODUCTION** |  |  |  |
| 2 | **Background** | Explain the scientific background and rationale for the reported study. What is the exposure? Is a potential causal relationship between exposure and outcome plausible? Justify why MR is a helpful method to address the study question | 3 | Molecular abnormalities detected in the blood that cause complex diseases represent an opportunity to identify biomarkers for both preventive and therapeutic interventions. These circulating biomolecules include proteins (enzymes and hormones), lipids, and metabolites that reflect physiological states of organ functions, immune response, and metabolism. Despite substantial observational evidence linking systemic biomarker levels with diverse health conditions, their causal relationships to complex diseases remain to be established, especially for diseases for which no reliable biomarkers exist. Previous studies that identify correlations often do not delineate cause and effect, thereby limiting the translational value of the findings.  The application of Mendelian Randomization (MR) represents an approach in addressing this critical gap. By utilizing genetic instruments as proxies for biomarker levels, MR, under specific assumptions, can control for confounding factors and reverse causation, offering insights into the causal effects of biomarkers on disease risks. However, the effects of many other biomarkers remain unclear, and existing MR studies have often been limited to specific biomarkers or disease categories. |
| 3 | **Objectives** | State specific objectives clearly, including pre-specified causal hypotheses (if any). State that MR is a method that, under specific assumptions, intends to estimate causal effects | 3 | Herein, we conducted a comprehensive MR analysis encompassing a broad spectrum of 212 biomarkers—including 115 circulating biomolecules measured through NMR3—and 99 human diseases to unveil previously obscured relationships of the circulating biomarkers’ role in disease etiology that may be leveraged for personalized prevention.  By utilizing genetic instruments as proxies for biomarker levels, MR, under specific assumptions, can control for confounding factors and reverse causation, offering insights into the causal effects of biomarkers on disease risks |
|  | **METHODS** |  |  |  |
| 4 | **Study design and data sources** | Present key elements of the study design early in the article. Consider including a table listing sources of data for all phases of the study. For each data source contributing to the analysis, describe the following: |  |  |
|  | a) | Setting: Describe the study design and the underlying population, if possible. Describe the setting, locations, and relevant dates, including periods of recruitment, exposure, follow-up, and data collection, when available. | 3-4 | MR-base offers access to multiple GWAS data including the MRC Integrative Epidemiology Unit (IEU) OpenGWAS database consisting of 26k GWAS reported in at least 379 different studies, covering ~40k individuals (in median) per study. Based on MRbase, we utilized all accessible exposure traits grouped under "Metabolites" category as well as other circulating biomarkers, and all available outcomes categorized as "Disease". All diseases we used are from datasets with IDs beginning with “ieu” (GWAS summary datasets generated by many consortia curated by IEU) and “bbj” (Biobank Japan), and the biomarkers are from “ukb” (UK Biobank analyses by IEU or Neale lab covering 28 biomarkers) and “met” (human blood biomarkers, immune markers, and circulating biomarkers analyzed by Shin et al. 2014 covering 76 biomarkers and Kettunen et al. 2016 covering 115 biomarkers). In instances where there are multiple summary datasets for the same trait (i.e., from different GWAS cohorts), we selected the top one from them based on the largest "sample size," "year," "number of SNPs," or "number of cases and controls," prioritizing the information available in this specified order. A list of all biomarkers used as exposures and information for their corresponding GWAS studies are available in Supplementary Table 1. Overall, we retained a total of 212 exposures (biomarkers) and 99 outcomes (diseases) for MR analyses. |
|  | b) | Participants: Give the eligibility criteria, and the sources and methods of selection of participants. Report the sample size, and whether any power or sample size calculations were carried out prior to the main analysis | 3-4 | See above. |
|  | c) | Describe measurement, quality control and selection of genetic variants | 3-4 | See above. |
|  | d) | For each exposure, outcome, and other relevant variables, describe methods of assessment and diagnostic criteria for diseases | 3-4 | See above. |
|  | e) | Provide details of ethics committee approval and participant informed consent, if relevant |  | N/A |
| 5 | **Assumptions** | Explicitly state the three core IV assumptions for the main analysis (relevance, independence and exclusion restriction) as well assumptions for any additional or sensitivity analysis | 4 | The three core assumptions for Mendelian Randomization (MR) analysis are:  Relevance: The genetic instruments (e.g., single nucleotide polymorphisms, SNPs) used as instrumental variables (IVs) should be associated with the exposure (circulating biomarkers in this case). This assumption ensures that the IVs are strong enough to influence the exposure. Herein, 2685 SNPs were found associated with 198 circulating biomarker exposures subject to the IV assumptions that were imposed by the standard two-sample MR approach (association threshold, P<5e-8; LD clumping cutoff r2>0.001 within 10Mb window) and were used for further analyses (Supplementary Table 2).  Independence: The genetic instruments should be independent of any confounding factors that may affect both the exposure and the outcome (disease). This assumption is justified by the random allocation of genetic variants. Further, each of the GWAS studies carefully control for covariates that may confound genetic associations. For example, in the UK Biobank biomarker GWAS conducted by the Neale lab, covariates include age, sex, age2, age*sex, age*sex2, and the first 20 PCs, making the final genetic instruments less prone to confounding. Additionally, the prior knowledge of demographic consistency can likely minimize the impact of unmeasured confounders due to similar demographic and genetic backgrounds.  Exclusion restriction: The genetic instruments should affect the outcome (disease) only through their effects on the exposure (circulating biomarkers) and not through any other direct or indirect pathways. This assumption ensures that the IVs influence the outcome solely via the exposure of interest. Critically, MR Egger was employed as a primary MR method in this study to detect and adjust for pleiotropy, where genetic variants may influence the outcome via pathways other than through the exposure.  To enhance robustness, we inferred causality using multiple MR methods, specifically inverse variance weighted, simple mode, weighted mode, weighted median, and MR Egger… … |
| 6 | **Statistical methods: main analysis** | Describe statistical methods and statistics used |  |  |
|  | a) | Describe how quantitative variables were handled in the analyses (i.e., scale, units, model) | 4 | For each method, the associations are reported with effect size and corresponding p-value statistics. The raw p-values are multi-testing corrected using the Bonferroni method. The confidence intervals (95%) are also calculated. Overall, robustness of the results is ensured based on the use of MR-Egger as discovery method, agreement across at least 4 MR methods, and Bonferroni correction. |
|  | b) | Describe how genetic variants were handled in the analyses and, if applicable, how their weights were selected | 4 | See Above |
|  | c) | Describe the MR estimator (e.g. two-stage least squares, Wald ratio) and related statistics. Detail the included covariates and, in case of two-sample MR, whether the same covariate set was used for adjustment in the two samples | 4 | See Above |
|  | d) | Explain how missing data were addressed |  | N/A |
|  | e) | If applicable, indicate how multiple testing was addressed | 4 | See Above |
| 7 | **Assessment of assumptions** | Describe any methods or prior knowledge used to assess the assumptions or justify their validity |  | See Section 5 above |
| 8 | **Sensitivity analyses and additional analyses** | Describe any sensitivity analyses or additional analyses performed (e.g. comparison of effect estimates from different approaches, independent replication, bias analytic techniques, validation of instruments, simulations) | 4 | See Section 5 Above |
| 9 | **Software and pre-registration** |  | 2 | This study is a two-sample MR study based on publicly available GWAS summary statistics data from MR-base resource (v0.5.6). The MR-base platform, developed by the MRC Integrative Epidemiology Unit at the University of Bristol, serves both as a database and an analytical framework accessed through the TwoSampleMR R package. |
|  | a) | Name statistical software and package(s), including version and settings used |  |  |
|  | b) | State whether the study protocol and details were pre-registered (as well as when and where) |  | N/A |
|  | **RESULTS** |  |  |  |
| 10 | **Descriptive data** |  |  |  |
|  | a) | Report the numbers of individuals at each stage of included studies and reasons for exclusion. Consider use of a flow diagram | 3-4 | See Section 4a Above |
|  | b) | Report summary statistics for phenotypic exposure(s), outcome(s), and other relevant variables (e.g. means, SDs, proportions) | 5 | A list of all tested relations, including IEU GWAS study IDs for each exposure/outcomes, and the summary statistics from MR analyses were included in Supplementary Table 2. |
|  | c) | If the data sources include meta-analyses of previous studies, provide the assessments of heterogeneity across these studies |  | N/A |
|  | d) | For two-sample MR:  i.  Provide justification of the similarity of the genetic variant-exposure associations between the exposure and outcome samples  ii.  Provide information on the number of individuals who overlap between the exposure and outcome studies |  | Given that this is a two-sample MR study, we provide two major justifications: (1) Similarity of the Genetic Variant-Exposure Associations Between the Exposure and Outcome Samples: Genetic instruments for the exposure were primarily derived from GWAS of circulating biomarkers conducted using data from the UK Biobank in addition to Shin and Kettunen et al., all of which predominantly consists of individuals of European ancestry. The outcome data were mainly sourced from case-control GWAS of European populations that likely share genetic architectures similar to those of the exposure study participants. (2) Number of Individuals Who Overlap Between the Exposure and Outcome Studies: Given the separate sources of the genetic data for exposures and outcomes (various case-control studies), and the timing of data availability and publication, it is reasonable to assume minimal overlap in the individual participants across these studies. Particularly, all significant results were found based on exposure (biomarker) GWAS from UK Biobank, which has its first genetic data published in 2018. The significant outcome GWAS for bipolar disorder, cardiovascular disease (CVD), myocardial infarction (MI), gout, and chronic kidney disease, all predated 2018. This temporal discrepancy between the data collection phases further supports the assumption of negligible overlap bias in our MR estimates. |
| 11 | **Main results** |  |  |  |
|  | a) | Report the associations between genetic variant and exposure, and between genetic variant and outcome, preferably on an interpretable scale | 5-6 | To ensure our results were robust, we focused on findings that had a significant (Bonferroni p-value < 0.05) association by MR-Egger and matched in 4 or more methods (with same effect direction and raw p-value < 0.05) based on MR-Egger’s robustness to directional pleiotropy relevant to circulating biomarkers. We identified a total 21 significant biomarker exposure vs. disease outcome associations (Figure 1). A list of all tested relations, including IEU GWAS study IDs for each exposure/outcomes, and the summary statistics from MR analyses were included in Supplementary Table 2. |
|  | b) | Report MR estimates of the relationship between exposure and outcome, and the measures of uncertainty from the MR analysis, on an interpretable scale, such as odds ratio or relative risk per SD difference | 7-8 | Figure 2 illustrates the 20 significant biomarker exposure-disease outcome associations with an effect size > \|0.1\|, p-value < 0.05, and agreement in effect directionality in 4 or more methods. The effect size cutoff > \|0.1\| was set to remove associations with marginal effect size to increase the likelihood of discovering more meaningful results.  Several biomarkers are associated with an increased risk of disease. The average effect size of total cholesterol and coronary heart disease is 0.66, ranging from 0.90 ± 0.22 (MR Egger) to 0.47 ± 0.13 (IVW). The association of total cholesterol and myocardial infarction has the average effect size of 0.61 with a range of (0.26, 1.13). For the apolipoprotein B and myocardial infarction, the average effect size is estimated as 0.59 with a range of (0.39, 0.90). For the glucose on bipolar disorder the average effect estimate is 0.39 with a range of (0.29, 0.55), while cystatin C has an inverse relationship on bipolar disorder with the average effect size of -0.31 ranging from -0.107 to -1 (Supplementary Table 1). |
|  | c) | If relevant, consider translating estimates of relative risk into absolute risk for a meaningful time period |  | N/A |
|  | d) | Consider plots to visualize results (e.g. forest plot, scatterplot of associations between genetic variants and outcome versus between genetic variants and exposure) | 5-8 |  |
| 12 | **Assessment of assumptions** |  |  |  |
|  | a) | Report the assessment of the validity of the assumptions |  | See Section 5 above |
|  | b) | Report any additional statistics (e.g., assessments of heterogeneity across genetic variants, such as *I^2^*, Q statistic or E-value) |  | See Section 5 above |
| 13 | **Sensitivity analyses and additional analyses** |  |  |  |
|  | a) | Report any sensitivity analyses to assess the robustness of the main results to violations of the assumptions | 5-8 | See 11a and 11b above |
|  | b) | Report results from other sensitivity analyses or additional analyses | 5-8 | See 11a and 11b above |
|  | c) | Report any assessment of direction of causal relationship (e.g., bidirectional MR) | 5-8 | See 11a and 11b above |
|  | d) | When relevant, report and compare with estimates from non-MR analyses | 5,7 | Of note, we discovered a strong, direct relationship between glucose and bipolar disorder and a strong, inverse relationship between cystatin C levels and bipolar disorder which, to the best of our knowledge, has never been directly reported before. Previous studies have demonstrated an association between impaired glucose metabolism and bipolar disorder in addition to increased prevalence of pre-diabetes and type 2 diabetes mellitus. Table 1 lists the identified exposure-outcome relationships along with previous supporting literature and evidence type. |
|  | e) | Consider additional plots to visualize results (e.g., leave-one-out analyses) |  |  |
|  | **DISCUSSION** |  |  |  |
| 14 | **Key results** | Summarize key results with reference to study objectives | 8 | We analyzed exposure-outcome relationships using 5 MR analysis methods to identify causal relationships between circulating biomarkers and diseases. In this study, significant results are defined as agreement between 4 or more analysis methods. Many of our demonstrated results replicated results from previous MR studies and other literature and none of our findings run contrary to previous literature. Additionally, our analysis also discovered two relationships of note: a direct relationship between glucose and bipolar disorder and an inverse relationship between cystatin C and bipolar disorder. |
| 15 | **Limitations** | Discuss limitations of the study, taking into account the validity of the IV assumptions, other sources of potential bias, and imprecision. Discuss both direction and magnitude of any potential bias and any efforts to address them | 9 | A limitation of this study is that much of the data comes from the UK biobank, a large-scale volunteer databank, which has a biased representation of Europeans and results may not generalize outside UK. However, UK biobank provides the largest GWAS datasets of most biomarkers studied herein and adequate statistical power; this bias may be overcome as other biobanks with diverse populations continue to expand. MR studies have their own limitations, including but not limited to: potential confounding factors, limitations of estimating associations for binary outcomes, pleiotropic effects, and population stratification. |
| 16 | **Interpretation** |  |  |  |
|  | a) | Meaning: Give a cautious overall interpretation of results in the context of their limitations and in comparison with other studies | 8-9 | Bipolar disorder is a neuropsychiatric disorder with multifactorial causes (genetic, trauma, exposure to certain medications, etc.) Our findings demonstrate two potential circulating markers that may contribute to development of bipolar disorder, high glucose and low cystatin C. Although high cystatin C levels have been linked with major depressive disorder, no studies, to the best of our knowledge, have demonstrated an association between cystatin C and bipolar disorder. Levels of Cystatin C, a natural inhibitor of cysteine proteases, are typically used clinically to assess kidney function. A common treatment for bipolar disorder is lithium, which has been shown to decrease renal function and can lead to elevated cystatin C levels, which may lend further insight to the mechanism behind lithium’s use as a treatment, however this remains to be confirmed. A connection between impaired glucose metabolism and bipolar disorder has been well documented in the literature, with over half of patients diagnosed with bipolar disorder also having insulin resistance, impaired glucose tolerance, or type 2 diabetes.7 Furthermore, it has been proposed that modulating the PI3K/Akt insulin signaling pathway may be a mechanism of lithium for the treatment of bipolar disorder.39 Therefore, our identification of a causal relationship between high glucose levels and bipolar disorder further supports the current body of evidence, and our findings for cystatin C highlight a novel finding that warrant investigation in subsequent studies. Our findings suggest that future studies aiming to reduce the risk of bipolar disorder could explore the lifestyle and clinical interventions that reduce blood glucose and maintain kidney function. |
|  | b) | Mechanism: Discuss underlying biological mechanisms that could drive a potential causal relationship between the investigated exposure and the outcome, and whether the gene-environment equivalence assumption is reasonable. Use causal language carefully, clarifying that IV estimates may provide causal effects only under certain assumptions | 8-9 | See Above. |
|  | c) | Clinical relevance: Discuss whether the results have clinical or public policy relevance, and to what extent they inform effect sizes of possible interventions | 8-9 | See Above. |
| 17 | **Generalizability** | Discuss the generalizability of the study results (a) to other populations, (b) across other exposure periods/timings, and (c) across other levels of exposure | 9 | Overall, by utilizing several MR methods as cross-validation, our study successfully reaffirmed several established connections in cardiovascular diseases, gout, and kidney diseases, while uncovering intriguing novel biomarkers that may be causal for bipolar disorder. These findings highlight the underappreciated roles that circulating biomarkers may play in disease mechanisms, which should open new avenues for targeted research and motivate development of precise diagnostic tools and therapeutic interventions. |
|  | **OTHER INFORMATION** |  |  |  |
| 18 | **Funding** | Describe sources of funding and the role of funders in the present study and, if applicable, sources of funding for the databases and original study or studies on which the present study is based | 10 | This work was supported in part through the computational and data resources and staff expertise provided by Scientific Computing and Data at the Icahn School of Medicine at Mount Sinai and supported by the Clinical and Translational Science Awards (CTSA) grant UL1TR004419 from the National Center for Advancing Translational Sciences. Research reported in this publication was also supported by the Office of Research Infrastructure of the National Institutes of Health under award number S10OD026880 and S10OD030463. The content is solely the responsibility of the authors and does not necessarily represent the official views of the National Institutes of Health. This work was supported by NIH NIGMS R35GM138113 and ACS RSG-22-115-01-DMC to KH and RF1AG072300 to JMC. |
| 19 | **Data and data sharing** | Provide the data used to perform all analyses or report where and how the data can be accessed, and reference these sources in the article. Provide the statistical code needed to reproduce the results in the article, or report whether the code is publicly accessible and if so, where | 10 | Data Availability  GWAS summary statistics data used can be found on MR-base resource (v0.5.6) through the ieu open GWAS project at the URL: https://gwas.mrcieu.ac.uk/.  Code Availability  The source code for the Mendelian randomization analyses is available at github.com/Huang-lab/MR. |
| 20 | **Conflicts of Interest** | All authors should declare all potential conflicts of interest | 10 | KH is a co-founder and board member of a non-for-profit organization, Open Box Science, where he does not receive any compensation. All other authors declare no competing interests. |

This checklist is copyrighted by the Equator Network under the Creative Commons Attribution 3.0 Unported (CC BY 3.0) license.

1.	Skrivankova VW, Richmond RC, Woolf BAR, Yarmolinsky J, Davies NM, Swanson SA, et al. Strengthening the Reporting of Observational Studies in Epidemiology using Mendelian Randomization (STROBE-MR) Statement. JAMA. 2021;under review.

2.	Skrivankova VW, Richmond RC, Woolf BAR, Davies NM, Swanson SA, VanderWeele TJ, et al. Strengthening the Reporting of Observational Studies in Epidemiology using Mendelian Randomisation (STROBE-MR): Explanation and Elaboration. BMJ. 2021;375:n2233.
